# Differential prediction performance between Caribbean- and Mainland-subgroups using state-of-the-art polygenic risk scores for coronary heart disease: Findings from the Hispanic Community Health Study/Study of Latinos (HCHS/SOL)

**DOI:** 10.1101/2024.09.25.24313663

**Authors:** Christina G. Hutten, Frederick J. Boehm, Jennifer A. Smith, Brian W. Spitzer, Sylvia Wassertheil-Smoller, Carmen R Isasi, Jianwen Cai, Jonathan T Unkart, Jiehuan Sun, Victoria Persky, Martha L Daviglus, Tamar Sofer, Maria Argos

## Abstract

**Background:** Coronary heart disease (CHD) is a leading cause of death for Hispanic/Latino populations in the United States. We evaluated polygenic risk scores (PRS) with incident myocardial infarction (MI) in a Hispanic/Latino study sample.

**Methods:** We leveraged data from the Hispanic Community Health Study/Study of Latinos (HCHS/SOL) to assess four CHD-PRS from the PGS catalog, derived using multiple methods (LDpred, AnnoPred, stacked clumping and thresholding, and LDPred2). We evaluated associations between each standardized PRS and time to adjudicated incident MI, adjusted for age, sex, first 5 principal components, and weighted for survey design. Concordance statistics (c-index) compared predictive accuracy of each PRS with, and in addition to, traditional risk factors (TRF) for CHD (obesity, hypercholesterolemia, hypertension, diabetes, and smoking). Analyses were stratified by self-reported Caribbean- (Puerto Rican, Dominican or Cuban) and Mainland- (those of Mexican, Central American, or South American) heritage subgroups.

**Results:** After 11 years follow-up, for 9055 participants (mean age (SD) 47.6(13.1), 62.2% female), the incidence of MI was 1.0% (n = 95). Each PRS was more strongly associated with MI among Mainland participants. LDPred2 + TRF performed best among the Mainland subgroup; HR=2.69, 95% CI [1.71, 4.20], c-index = 0.897, 95% CI [0.848, 0.946]; a modest increase over TRF alone, c-index = 0.880, 95% CI [0.827, 0.933]. AnnoPred + TRF performed best among the Caribbean sample; c-index = 0.721, 95% CI [0.647, 0.795]; however, was not significantly associated with rate of MI (HR=1.14, 95% CI [0.82, 1.60]).

**Conclusion:** PRS performance for CHD is lacking for Hispanics/Latinos of Caribbean origin who have substantial proportions of African genetic ancestry, risking increased health disparities. AnnoPred, using functional annotations, outperformed other PRS in the Caribbean subgroup, suggesting a potential strategy for PRS construction in diverse populations. These results underscore the need to optimize cumulative genetic risk prediction of CHD in diverse Hispanic/Latino populations.

## Background

About 20.5 million Americans have coronary heart disease (CHD) and 720,000 will have a new coronary event this year (1). The rates of CHD in the Hispanic/Latino communities are similar to the non-Hispanic White population; however, risk factors for CHD are more prevalent among Hispanics/Latinos (2). Projections estimate Hispanic/Latino populations will represent 28% of the U.S. population by 2060 (3). Thus, tools to identify high-risk individuals are paramount to initiate preventive measures and mitigate CHD morbidity and mortality for Hispanic/Latino populations.

Precision medicine promises to use genetic information to target individuals with elevated disease risk and personalize treatments. Polygenic risk scores (PRS) are weighted or non-weighted sums of risk-conferring alleles of single nucleotide polymorphisms (SNPs) and may improve risk prediction over traditional risk factors (TRF) alone (4–8). A major limitation of the existing genetic epidemiology literature is a lack of diversity in study samples which limits generalizability of findings and can contribute to disparities in healthcare and personalized medicine for underrepresented populations (9).

Hispanic/Latino populations living in the U.S. are highly diverse, admixed populations represented by varied genetic ancestries (European, African, and/or Amerindian), as well as varied cultures and environmental exposures (10). Given this genetic diversity, performance of PRS developed using SNPs associated with CHD in European ancestry populations is underwhelming due to differences in linkage disequilibrium (LD), allele frequencies and effect sizes (11). In a large cohort of Hispanics/Latinos in the U.S., we assessed the ability of four CHD PRS, derived using varying methods, to predict incident myocardial infarction (MI) and determine whether prediction is improved over traditional CHD risk factors.

## Methods

### Study Population

The Hispanic Community Health Study/Study of Latinos (HCHS/SOL) is a large cohort of Hispanic/Latino health, comprising 16,415 participants aged 18-74 years. As a multicenter-epidemiologic study to evaluate and identify risk and protective factors with the health of U.S. Hispanics/Latinos, recruitment was conducted using a two-stage area probability sampling of households in Chicago, San Diego, Bronx, and Miami, and enrollment occurred at one of four field centers in each location. (12,13). Institutional Review Board (IRB) approval was obtained at each center’s respective IRB, and participants provided written informed consent in their preferred language (English or Spanish). Participants underwent an extensive clinical exam and assessments at baseline (Visit 1: 2008-2011) and follow-up (Visit 2: 2015-2017). Additional telephone follow-up continued through 2019.

Of the 16,415 HCHS/SOL participants, 11,623 returned for their Visit 2 exam, and 11,469 provided consent at the Visit 2 examination for continued use of their DNA samples in genetic research by HCHS/SOL affiliated investigators. Of those who provided consent for the use of genetic data and for whom complete Visit 1 and Visit 2 data were available on key covariates were included in the current analyses (n=9055). Those without genotype data (n = 1807) were omitted from PRS analyses (**Supplemental Figure 1**).

### Clinical evaluations in the HCHS/SOL

Visit 1 and 2 examinations were conducted by trained/certified health interviewers at each field center according to standard protocols (14). Participants were asked to fast and abstain from smoking 12 hours and avoid vigorous physical activity on the morning of the examination. Anthropometric characteristics were measured, and body mass index (BMI) was calculated as weight in kilograms divided by height in meters squared(15). Three seated blood pressure measurements were obtained after a 5-minute rest; the average of the second and third was calculated for use in analyses (12,15).

### Medication use in the HCHS/SOL

All prescription and over-the-counter medications used in the four weeks leading up to the Visit 1 examination were ascertained via two methods: 1) participants brought all medication containers to the interview where they were recorded, and 2) participants self-reported which medications were for specific conditions, including high blood pressure and diabetes. Antihypertensive, antidiabetic, and lipid-modifying medication use was defined as either transcribed or self-reported using the Master Drug Data Base (Medispan MDDB®).

### Laboratory evaluation in the HCHS/SOL

Fasting blood samples were shipped to the HCHS/SOL Central Laboratory at the University of Minnesota and measured for: total cholesterol using a cholesterol oxidase enzymatic method; high-density lipoprotein (HDL) cholesterol using a direct magnesium/dextran sulfate method; plasma glucose using a hexokinase enzymatic method; serum triglycerides using a glycerol blanking enzymatic method (Roche Diagnostics, Indianapolis, IN); low-density lipoprotein (LDL) cholesterol was calculated using the Friedewald equation (16); Hemoglobin A1c (HbA1c) was measured using a Tosoh G7 Automated HPLC Analyzer (Tosoh Bioscience) (15).

### Outcomes

Incident MI was based on participant-reported hospitalization or emergency room (ER) visit during annual follow-up phone interview or at the Visit 2 exam. Medical records review of hospital and ER visits for MI events were abstracted and adjudicated. First incident MI events were reviewed by 2 independent reviewers, with discrepancies settled by an adjudicator. Follow-up time to first MI event was defined as the difference between the date of the first MI event and the Visit 1 exam date. If no MI event occurred, follow-up time was determined by censor date (date of death or date of withdrawal) or date of last follow-up.

### Genotyping and Imputation

HCHS/SOL participants who consented to genetic studies at Visit 1 had DNA extracted from whole blood samples and genotyped using a customized HCHS/SOL Illumina Omni 2.5 M array (HumanOmni2.5-8 v.1-1) (17–19). Standard quality assurance and quality control measures were applied to generate recommended variant- and sample-level quality filters (19,20). There were 2,232,944 genetic variants that passed quality filters and were informative that proceeded for imputation (10). Genome-wide imputation was performed via the Michigan imputation server using the TOPMed 2.0 imputation panel (21,22). Imputation quality was reported for each variant (R^2^).

### Polygenic Risk Scores

The PRS were selected from the PGS catalog (23) to analyze several PRS with varying numbers of SNPs, methods for construction, and genome-wide association (GWAS) discovery populations. Summary statistics were downloaded from the PGS catalog(23). Only variants with imputation quality R^2^ ≥ 0.8 and minor allele frequency ≥0.01 were used. PRSs were constructed from summary statistics using the PRSice software (24), without any clumping and thresholding. The scores were standardized to mean zero and variance one in the analytic sample. The four PRS are summarized in **Table 1** and methodology for construction is summarized below:

a. <u>PGS000013</u> (25)<u>-LDpred</u> (26): Bayesian approach used to calculate posterior mean effect size for each SNP based on prior GWAS effect sizes and modeled LD information from an external reference population (25,26).
b. <u>PGS001355</u> (27)<u>-AnnoPred</u> (28): Used functional annotations to estimate prior SNP effect sizes, incorporated in a Bayesian framework and jointly modeled with an estimated LD matrix from reference genotype panels and inferred posterior SNP effect sizes (27,28).
c. <u>PGS002776</u> (29)<u>-SCT</u> (30): Stacked clumping and thresholding (SCT) first set a clumping window (*kb*), correlation (r^2^) and p-value thresholds to select SNPs into a PRS. A set of parameters is chosen for LD, window size, p-value, and INFO score (based on quality of imputation) (30). Clumping and thresholding are then run on each combination of these parameters using the R package ‘bigsnpr’ (31) to provide a PRS for each combination. Using penalized regression modeling, the PRS are stacked to produce a set of weights to apply to each SNP in prediction modeling (29,30).
d. <u>PGS003725</u> (32)<u>-LDPred2</u> (33): Bayesian approach to calculate a posterior mean effect size for each SNP based on prior GWAS effect sizes followed by shrinkage using LD information (32,33).

**Table 1.** Characteristics of PRS selected from the PGS Catalog ^(23)^

| PRS | Method | Number of SNPs | GWAS population | Training population | Evaluation population | Reference |
| --- | --- | --- | --- | --- | --- | --- |
| PGS000013 | LDPred | 6,630,150 | Multi-ancestry(75.3% European, 13.6% South Asian, 6% East Asian, 2.2% Hispanic or Latin American, 1.7% African, 1.2% Greater Middle Eastern) | 100% European | Multi-ancestry (49.2% European, 15.9% Multi-ancestry (including European), 9.5% African, 9.5% Hispanic or Latin American, 6.3% South Asian, 4.8% East Asian, 3.2% Not Reported, 1.6% Additional Asian Ancestries) | PMID: 30104762 |
| PGS001355 | AnnoPred | 2,994,055 | 100% European | 100% European | 100% European | PMID: 33433237 |
| PGS002776 | SCT | 390,782 | Multi-ancestry (75.3% European, 13.6% South Asian, 6% East Asian, 2.2% Hispanic or Latin American, 1.7% African, 1.2% Greater Middle Eastern) | 100% European | 100% European | PMID: 36459520 |
| PGS003725 | LDpred2 | 1,296,272 | Multi-ancestry (76.4% European, 5.3% African, 14.7% East Asian, 2.1% Hispanic of Latin American, 1.5% South Asian) | 100% European | Multi-ancestry (25% African, 25% European, 25% South Asian, 12.5% Hispanic or Latin American, 12.5% East Asian) | PMID: 37414900 |

Note: Table provides PRS summary data based on information available in the PGS Catalog repository (23) or the respective manuscript.

### Traditional risk factors

Traditional risk factors (TRF) were evaluated in comparison to and in conjunction with each PRS for predictiveness and defined as follows: Hypercholesterolemia (total cholesterol of ≥240mg/dL, LDL cholesterol ≥160mg/dL, HDL <40mg/dL, or receiving cholesterol-lowering medication); hypertension (systolic blood pressure ≥140mmHg, diastolic blood pressure ≥90mmHg, or use of high blood pressure medication); hypertension AHA (systolic blood pressure ≥130mmHg, diastolic blood pressure ≥80mmHg based on the 2017 ACC/AHA Guidelines definition, or use of high blood pressure medication (34); obesity (body mass index ≥30kg/m^2^ at Visit 1); diabetes mellitus (fasting plasma glucose ≥126mg/dL, 2-hour post-load plasma glucose ≥200mg/dL, HbA1c ≥6.5%, or use of antihyperglycemic medications); and smoking (self-reported current cigarette smoking) (15).

### Statistical Analysis

All reported values were weighted to adjust for complex survey design, sampling probability, and non-response in the HCHS/SOL cohort. The calculation of the sampling weights for Visit 2 was based on the sampling weights for Visit 1 and accounted for the participant non-response for Visit 2. Chi-square tests were used to test for significant differences in baseline characteristics and incident MI.

Each PRS was modeled continuously. Multivariable Cox proportional hazards models were used to assess the association of each standardized PRS adjusted by *a priori* confounders: age, sex, and the first 5 principal components (PCs) to account for genetic ancestry and population structure. PC analysis was performed previously (detailed methods in reference 12), which showed no further benefit to controlling for confounding by ancestry beyond 5 PCs (10). Statistical evaluation of interaction by sex was conducted. We also assessed the associations between each PRS with incident MI stratified by self-reported Caribbean-(Puerto Rican, Dominican, or Cuban heritage) and Mainland- (Mexican, Central American, or South American heritage) Hispanic/Latino subgroups using Cox proportional hazards regression and adjusted for age, sex and the first 5 PCs. Sensitivity analyses were conducted to assess associations of each PRS with incident MI when restricted to participants 50 years and older while stratified by Caribbean- and Mainland-subgroups.

To determine whether the addition of each PRS improves the prediction of incident MI beyond TRF (hypertension, high cholesterol, diabetes, obesity, and smoking) we used the concordance statistic (c-index) (35). The c-index was calculated for each of the TRF alone, each PRS alone, the TRF combined, and for each PRS+TRF combined.

## Results

For the analytic sample (n = 9055), mean age was 47.6 years (SD: 13.1), 62.2% were female, with 1% incidence of MI (n = 95) over a median 9.8 years of follow-up (IQR: 9.1-10.6 years) **(Table 2, Supplemental Figure 1)**. In unadjusted analysis, increased risk of incident MI was associated with age, Cuban background, Caribbean origin, less than- or greater than- a high school degree or GED, hypertension, diabetes mellitus, and current smoking status (**Table 2**). Study participation with the San Diego field center was associated with lower risk of incident MI. Each standardized PRS was normally distributed (**Figure 1**). When stratified by Mainland and Caribbean subgroups, the SCT PRS for Mainland subgroup showed a higher median (IQR) than Caribbean subgroup while the LDPred2 PRS elicited a higher median (IQR) distribution for the Caribbean subgroup (**Supplemental Figure 2**). Baseline characteristics of the Mainland versus Caribbean subgroups are presented in **Supplemental Table 1**.

**Figure 1.**
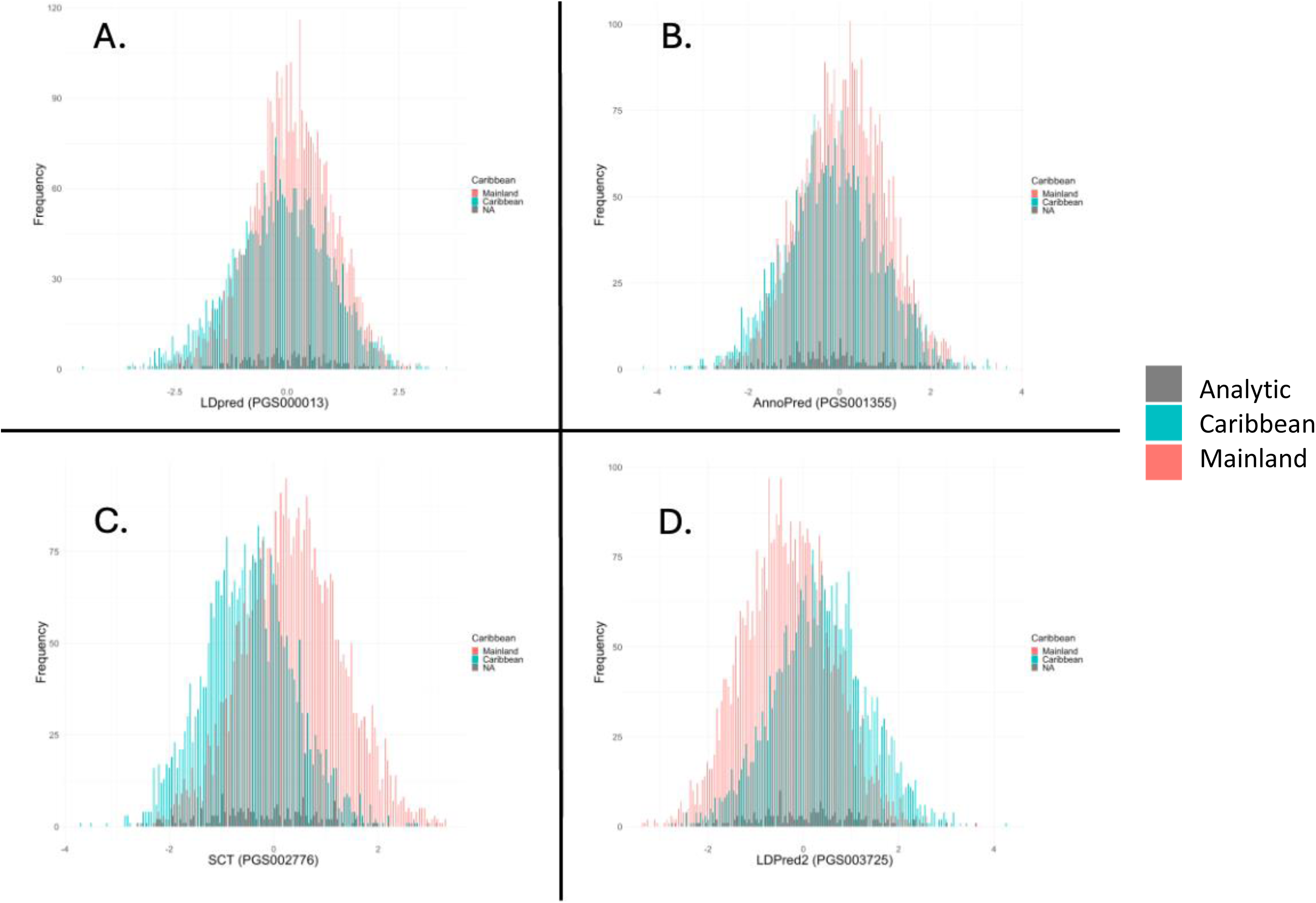
Standardized PRS distributions stratified by Caribbean and Mainland subgroups. A. LDPred, B. AnnoPred, C. SCT, D. LDPred2. Blue = Caribbean subgroup, Red = Mainland subgroup

**Table 2.** Baseline characteristics in relation to adjudicated incident myocardial infarction through 2019.

|  | n | Incident MI |  |
| --- | --- | --- | --- |
|  |  | Number of events | HR (95% CI) |
| **Sample baseline characteristics** | 9055 | 95 |  |
| Sex |  |  | P <0.001 |
| Males | 3421 | 56 | Reference |
| Females | 5634 | 39 | 0.63 (0.33, 1.21) |
| Age (years) |  |  | P <0.001 |
| 18-39 | 2244 | 6 | Reference |
| 40-49 | 2470 | 21 | 1.25 (0.39, 4.00) |
| 50-59 | 2676 | 46 | **4.22 (1.40, 12.73)** |
| 60+ | 1665 | 22 | 3.15 (0.98, 10.06) |
| Hispanic/Latino background |  |  | P = 0.4 |
| Mexican | 3515 | 29 | Reference |
| Central American | 942 | 8 | 0.95 (0.39, 2.31) |
| Cuban | 1426 | 22 | **2.14 (1.15, 3.97)** |
| Dominican | 839 | 8 | 2.93 (0.84, 10.22) |
| Puerto Rican | 1467 | 18 | 1.43 (0.68, 2.98) |
| South American | 618 | 6 | 1.80 (0.60, 5.37) |
| More than one/other heritage/NA | 248 | 4 | -- |
| Background Strata |  |  | P = 0.1 |
| Mainland | 5075 | 43 | Reference |
| Caribbean | 3732 | 48 | **1.92 (1.09, 3.37)** |
| More than one/other heritage/NA | 248 | 4 | -- |
| Study Center |  |  | P = 0.1 |
| Bronx | 2157 | 21 | Reference |
| Chicago | 2282 | 25 | 0.89 (0.36, 2.20) |
| Miami | 2402 | 34 | 0.96 (0.41, 2.22) |
| San Diego | 2214 | 15 | **0.35 (0.14, 0.88)** |
| Education |  |  | P = 0.044 |
| No high school diploma or GED | 3335 | 42 | **2.19 (1.18, 4.09)** |
| At most a High School diploma or GED | 2279 | 22 | Ref |
| Greater than High school or GED | 3428 | 30 | **2.14 (1.01, 4.54)** |
| Health insurance |  |  | P = 0.1 |
| Does not have health insurance | 4288 | 43 | Ref |
| Has health insurance | 4675 | 49 | 1.41 (0.77, 2.57) |
| Total Physical activity levels |  |  | P = 0.2 |
| High | 875 | 15 | Ref |
| Moderate | 4004 | 35 | 0.51 (0.19, 1.38) |
| Low | 4150 | 45 | 0.64 (0.26, 1.56) |
| Lipid Lowering Medications | 1236 | 22 | 1.76 (0.95, 3.277) |
| Statin users | 1135 | 19 | 1.64 (0.85, 3.14) |
| **CHD risk factors at Visit 1** |  |  |  |
| High total cholesterol | 4243 | 66 | 1.60 (0.82, 3.14) |
| Dyslipidemia | 3605 | 54 | 1.35 (0.73, 2.48) |
| Hypertension (>140/90) | 2653 | 54 | **3.34 (1.79, 6.24)** |
| AHA updated 2017 Hypertension | 4236 | 75 | **2.97 (1.33, 6.63)** |
| (>130/80) |  |  |  |
| Obesity ($\geq 30 \text{ kg/m}^2$)<br>(ref = $18.5\text{-}25 \text{ kg/m}^2$) | 3897 | 36 | 1.06 (0.34, 3.33) |
| Diabetes Mellitus | 1970 | 39 | **3.93 (1.43, 10.82)** |
| Current Smoker | 1664 | 34 | **2.24 (1.16, 4.33)** |

*Note:* All values (except N) are weighted for study design and non-response.

Multivariable-adjusted associations of each standardized-PRS with incident MI are presented in **Figure 2**. For every one-standard deviation (SD) increase in LDPred2 PRS, the Mainland subgroup had 2.69 [95% CI, 1.72-4.20] higher risk of incident MI while the Caribbean group showed no increased risk (HR 1.01 [95% CI, 0.65-1.56]). Similarly, the LDPred PRS had 2-times higher risk of incident MI in the Mainland subgroup (HR 1.97 [95% CI, 1.23-3.15]) with every one-SD increase in PRS; however, while risk increased for the Caribbean subgroup, it was not significant (HR 1.15 [95% CI, 0.87-1.51]). The AnnoPred PRS showed 48% higher risk of MI [95% CI, 1.15-1.91] with every one-SD increase in PRS; however, when stratified by subgroup, the Mainland group showed 80% higher risk of incident MI [95% CI, 1.20-2.72] and Caribbean group had no increased risk. The SCT PRS demarcated no significantly increased risk for any subgroup (**Figure 2**). Sensitivity analysis for participants over 50 years remained consistent regarding magnitude and significance of the associations for each PRS stratified by Caribbean and Mainland groups (**Supplemental Table 2**). There was no evidence of heterogeneity of effects by sex for LDPred2 and SCT PRS (interaction p values = 0.17 and 0.52, respectively) while there was a significant interaction by sex for LDPred and AnnoPred PRS (interaction p values = 0.04 and 0.03, respectively) where higher risk was observed among females (**Supplemental Table 3)**.

**Figure 2.**
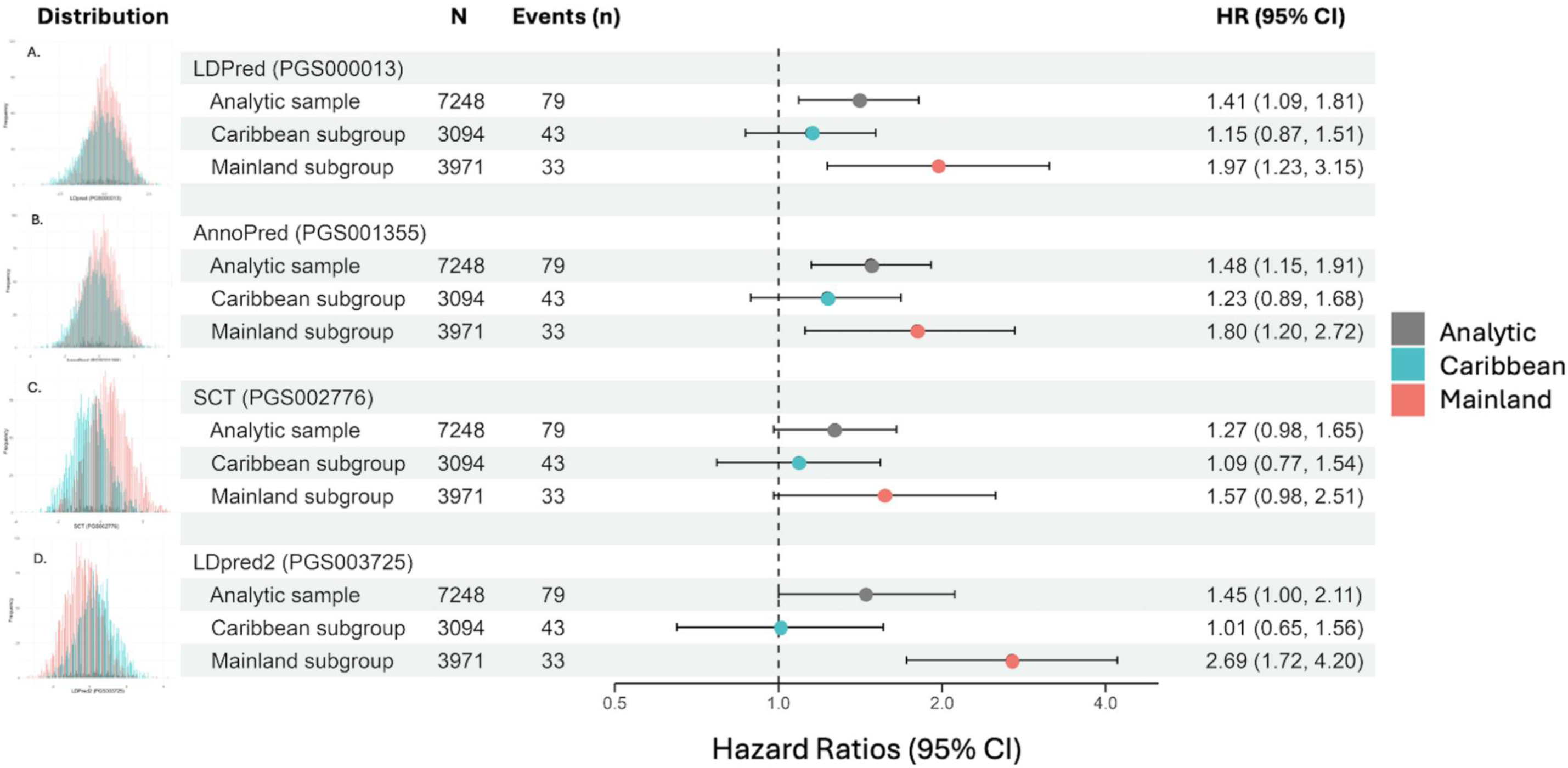
Cox proportional hazards regression model associations of each standardized PRS with incident MI outcomes stratified by Caribbean and Mainland subgroups. A. LDPred, B. AnnoPred, C. SCT, D. LDPred2. Blue = Caribbean subgroup, Red = Mainland subgroup. Models were adjusted for age, sex, the first 5 principal components, and weighted for complex survey design.

To evaluate predictive probability of traditional risk factors (TRF) in comparison to each PRS, we used Cox proportional hazards regression to model the 5 TRF separately (BMI, high total cholesterol, hypertension, diabetes, and smoking), the 5 TRF together, and the 5 TRF together with each PRS. Each model was adjusted for age, sex, the first 5 PCs, and weighted for complex survey design. Each PRS, TRF, and PRS+TRF performed best at predicting incident MI in the Mainland strata (c-index range: 0.809-0.897); highest for the model that included LDPred2+TRF (c-index: 0.897, SE: 0.025) (**Figure 3**) and an improvement of 0.017 over prediction by combined TRF. The SCT+TRF performed worse than TRF combined for the Mainland subgroup while AnnoPred+TRF (c-index: 0.883, SE: 0.029) and LDPred+TRF (c-index: 0.884, SE: 0.029) each provided slight improvement. Each PRS alone performed worse in the Mainland subgroup than TRF combined. LDPred2 alone predicted incident MI better than BMI, high total cholesterol or smoking alone.

**Figure 3.**
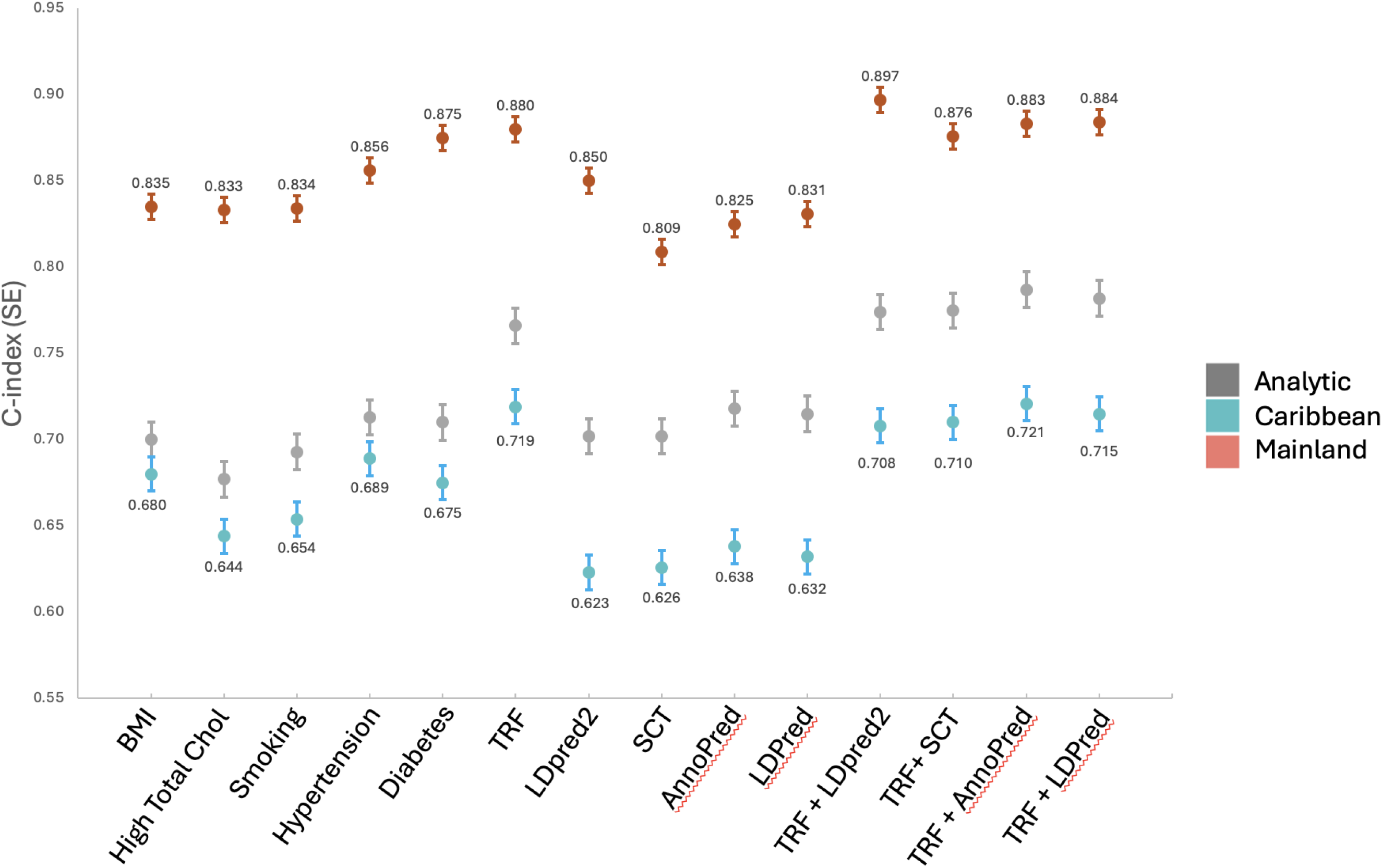
Concordance statistic (C-index). Cox proportional hazards regression models for the associations between each PRS and incident MI for traditional risk factors individually and in combination with each PRS. All models were adjusted for age, sex, and the first 5 principal components. TRF = Traditional risk factors; BMI = body mass index; High Total Chol = High Total Cholesterol; Smoking = current smoking status; Analytic (Gray) = full analytic sample; Caribbean (Blue) = self-reported Cuban, Dominican Republic, and Puerto Rican heritage; Mainland (Red) = self-reported Mexican, Central American, and South American heritage groups.

The AnnoPred PRS+TRF performed best in the Caribbean subgroup (c-index: 0.721, SE: 0.038), an improvement of only 0.002 over the combined TRF. Ever other PRS+TRF combination decreased prediction of incident MI for the Caribbean subgroup below TRF combined. Each PRS alone performed worse than each separate TRF. The AnnoPred PRS+TRF also performed best in the full analytic sample (c-index: 0.787, SE: 0.036) which improved prediction 0.021 over TRF combined. TRF combined performed better than each PRS alone by 0.048-0.064 increase in c-index for the analytic sample, while each PRS+TRF also improved performance slightly over the combined TRF (**Figure 3**).

## Discussion

In the current study, we utilized four comprehensive PRS associated with CHD to assess their prediction of incident MI in a diverse cohort of Hispanics/Latinos in the U.S. Overall, each PRS predicted incident MI best for the Mainland subgroup. AnnoPred PRS had improved performance for the full analytic sample and Caribbean strata over other PRS, which suggests improved utility among those with heritage from Cuba, the Dominican Republic, and Puerto Rico. This may indicate a potential avenue for methods development in PRS construction to improve prediction in African-admixed populations.

Incorporating genetic information into risk prediction tools improves performance. Inouye et al. (2018) compared the predictive value of TRF alone, TRF combined, and PRS+TRF for risk prediction of CHD in the UK Biobank, a cohort of primarily European ancestry. Similar to our findings, each TRF by itself (smoking, diabetes, family history of heart disease, body mass index, hypertension, and high cholesterol) did not perform as well as the PRS at predicting CHD and PRS+TRF showed the best predictive value for CHD by C-index (36). We also found each TRF alone had slightly lower predictive value than 5 TRF combined. The PRS+TRF had even higher predictive value in some instances, such as LDPred+TRF for the Mainland subgroup. While AnnoPred+TRF also showed higher predictive value for the full analytic sample and Caribbean strata, c-index improvement was only modest in all groups. This suggests some PRS may be useful for CHD risk prediction in subgroups of Hispanics/Latinos early in life, before TRF develop.

Comparing relative risk for CHD using TRF (e.g., cholesterol, smoking status, and systolic blood pressure) versus PRS+TRF could lead someone to take preventive measures earlier (37). Given the relatively young age of Hispanic/Latino populations in the U.S. (38), identifying those at increased genetic risk may lessen the burden of CHD events by identifying those in need of primary prevention rather than rely on current clinical guidelines which only incorporate TRF (15,39,40). We found the predictive value of LDPred2+TRF to perform better than TRF combined and suggests the use of a PRS provides an ideal opportunity for preventive management.

Hispanic/Latino populations are highly admixed populations with ancestry influenced by European, African, and Amerindian backgrounds (10). Our analysis shows evidence of PRS prediction differences between strata of Mainland and Caribbean subgroups. The Mainland subgroup, with heritage from Mexico, South America, and Central America, tends to include individuals with equal proportions of European and Amerindian genetic ancestry and a lower proportion of African ancestry (10). Alternatively, the Caribbean subgroup tends to consist of individuals with a large proportion of European and African ancestries and a lower proportion of Amerindian ancestry (10). Despite the large proportion of European admixture, each PRS performed worse in the Caribbean subgroup. Previous principal components analysis of Caribbean Hispanic/Latino individuals traced genetic ancestry to Spain and Portugal; however, the distance of genetic ancestry from elsewhere in Europe suggests a bottleneck and genetic drift that occurred when Europeans settled in the Caribbean (41). Each GWAS used for construction of PRS, may not include variants in LD with African populations and may not have sampled participants from the Iberian peninsula. Interestingly, the PRS that performed best in the Caribbean subgroup was the AnnoPred, which was developed, trained, and evaluated in European cohorts (27). Another study using data from the Million Veteran Program identified heterogeneity in PRS validity among Hispanics when stratified by self-identified race/ethnicity principal components (42). Our analysis provides further support that PRS use should consider Hispanic/Latino populations as distinct groups.

The portability of PRS between populations has come into question due to differences in LD, allele frequencies, and genetic architecture (9,43); however, we hypothesized a more diverse sample of Hispanics/Latinos, such as HCHS/SOL, would provide a higher likelihood that the SNPs are in LD with a causal variant. This may be why each PRS conferred increased risk for incident MI in the Mainland subgroup. Previous work has shown selecting genetic variants from the robust GWAS literature in European ancestry populations generally performs well in Hispanic/Latino populations (44). The LDPred and LDPred2 PRS both utilized multi-ancestry GWAS for SNP selection and evaluation (25,32). The additional step used in LDPred2 using shrinkage by LD may have improved its performance, although only in the Mainland group.

Furthermore, we provide evidence that a larger number of SNPs does not always lead to improved performance and may differ by genetic ancestry. LDPred2 contained 5 million less SNPs than the original LDPred and while using similar Bayesian methods for construction, LDPred2 conferred higher risk of incident MI for every 1-SD increase in PRS compared to LDPred for the Mainland sample. Consistent with our findings, the eMERGE network assessed a 1.7 million SNP PRS for incident CHD compared to the same LDPred PRS utilized here with 6.6 million SNPs in a self-reported Hispanic sample of 2500 individuals. The 1.7 million PRS performed better than the larger LDPred PRS according to c-index (0.683 vs. 0.659, respectively), despite having fewer SNPs included (45). However, LDPred PRS performed better in the analytic and Caribbean subset for HCHS/SOL, which may have benefited from the additional 5 million SNPs providing a higher chance that those included were in LD. Similarly, AnnoPred contains nearly 2 million more SNPs than LDPred2 and was the best performing in the Caribbean subgroup.

Our findings extend the understanding of genetic contributions to CHD in Hispanic/Latino populations and, thus, prevent expanding health disparities as we enter the era of precision medicine. Most genetic research has been conducted in populations with overwhelmingly high percentages of European genetic ancestry (9). From our analysis, it is apparent that genetic ancestry plays a role in predicting incident MI with PRS. More accurate predictions may be possible by considering European, African, and/or Amerindian ancestry proportions. The PRS assessed in this study may not be the most predictive tool for use in Hispanic/Latino populations; however, it is promising the PRS were associated with increased risk of incident MI and that some associations were more pronounced in certain strata. Identifying additional SNP-CHD associations in Hispanic/Latino populations may improve PRS-based CHD predictions for these populations.

The present study has several strengths. This is one of the first studies to provide insight into the genetic contribution to CHD for Hispanic/Latino populations in the U.S. using one of the largest and most diverse prospective longitudinal studies of Hispanic/Latino health in the U.S. We had access to well-characterized baseline and follow-up data, including genotype data. Despite the large and diverse cohort of Hispanics/Latinos, study participants were relatively young, with an average age of 41.6 years at Visit 1. Given subjects’ young ages, we accrued a relatively small number of CHD events. However, despite the low event count, we identified several significant PRS-CHD associations. Further, the definition of CHD used to create each PRS may differ from our outcome definition, which only included incident MI. However, each event was adjudicated, lowering the likelihood of misclassification.

Utilization of a PRS may help ameliorate the burden of CHD for Hispanic/Latino populations in the U.S. by identifying high-risk individuals for implementing preventive measures at an earlier timepoint than is possible when using traditional risk factors (TRF) alone. The LDPred2 PRS shows promise in predicting CHD events in Mainland Hispanic/Latino populations originating from Mexico, Central America, and South America, while AnnoPred PRS shows promise as a method for PRS development to improve risk prediction in Caribbean Hispanics/Latinos with Cuban, Dominican and Puerto Rican ancestry. Future research with a greater number of CHD events will provide further evidence for the utility of PRS in Hispanic/Latino populations in the U.S.

## Data Availability

Requests for data can be addressed to the

---

**Supplementary Table 1.** Baseline characteristics by Caribbean vs. Mainland subgroups.

|  |  | Caribbean | Caribbean<br>No. of Incident<br>MI events | Mainland | Mainland<br>No. of Incident<br>MI events | P-value |
| --- | --- | --- | --- | --- | --- | --- |
|  | N | n (%) | n | n (%) |  |  |
| Total | 8807 | 3732 (42.4) | 48 | 5075 (57.6) | 43 |  |
| Sex |  |  |  |  |  | P<br><0.015 |
| Females | 5634 | 2274 (60.9) | 22 | 3222 (63.5) | 15 |  |
| Age (years) |  |  |  |  |  | P<br><0.001 |
| 18-39 | 2244 | 689 (18.7) | 3 | 1451 (28.6) | 2 |  |
| 40-49 | 2470 | 1004 (26.9) | 9 | 1398 (27.5) | 11 |  |
| 50-59 | 2676 | 1173 (31.4) | 22 | 1447 (28.5) | 22 |  |
| 60+ | 1665 | 857 (23.0) | 14 | 779 (15.3) | 8 |  |
| Hispanic/Latino background |  |  |  |  |  | NA |
| Mexican | 3515 | 0 (0.0) | NA | 3515 (69.3%) | 29 |  |
| Central American | 942 | 0 (0.0) | NA | 942 (18.6%) | 8 |  |
| Cuban | 1426 | 1426 (38.2) | 22 | 0 (0.0) | NA |  |
| Dominican | 839 | 839 (22.5) | 8 | 0 (0.0) | NA |  |
| Puerto Rican | 1467 | 1467 (39.3) | 18 | 0 (0.0) | NA |  |
| South American | 618 | 0 (0.0) | NA | 618 (12.2) | 6 |  |
| Study Center |  |  |  |  |  | <0.001 |
| Bronx | 2157 | 1753 (47.0) | 17 | 315 (6.2) | 1 |  |
| Chicago | 2282 | 474 (12.7) | 8 | 1754 (34.6) | 17 |  |
| Miami | 2402 | 1479 (39.6) | 23 | 870 (17.1) | 11 |  |
| San Diego | 2214 | 26 (0.7) | 0 | 2136 (42.1) | 14 |  |
| Education |  |  |  |  |  | <0.001 |
| No high school diploma or GED | 3335 | 1246 (33.4) | 19 | 2019 (39.8) | 21 |  |
| At most a High School diploma or GED | 2279 | 962 (25.8) | 13 | 1274 (25.1) | 9 |  |
| Greater than High school or GED | 3428 | 1524 (40.8) | 16 | 1776 (35.0) | 12 |  |
| Health insurance |  |  |  |  |  | <0.001 |
| Does not have health insurance | 4288 | 1307 (35.0) | 18 | 2878 (56.7) | 24 |  |
| Has health insurance | 4675 | 2367 (63.4) | 28 | 2166 (42.7) | 18 |  |
| Total Physical activity levels |  |  |  |  |  | <0.001 |
| High | 875 | 314 (8.4) | 7 | 521 (10.3) | 7 |  |
| Moderate | 4004 | 1568 (42.0) | 17 | 2324 (45.8) | 18 |  |
| Low | 4150 | 1843 (49.4) | 24 | 2217 (43.7) | 18 |  |
| Lipid Lowering Medications | 1236 | 643 (17.2) | 13 | 568 (11.2) | 9 | <0.001 |
| Statin users | 1135 | 512 (10.1) | 13 | 599 (16.1) | 6 | <0.001 |
| **CHD risk factors at Visit 1** |  |  |  |  |  |  |
| High total cholesterol | 4243 | 1839 (49.3) | 35 | 2305 (45.4) | 28 | <0.001 |
| Dyslipidemia | 3605 | 1476 (39.5) | 28 | 2043 (40.3) | 23 | 0.348 |
| Hypertension (>140/90) | 2653 | 1426 (38.2) | 27 | 1161 (22.9) | 27 | <0.001 |
| AHA updated 2017 Hypertension (>130/80) | 4236 | 2183 (58.5) | 38 | 1938 (38.2) | 35 | <0.001 |
| Obesity ($\geq 30\text{kg/m}^2$) | 3897 | 1624 (43.5) | 19 | 2144 (42.2) | 17 | 0.510 |
| Diabetes Mellitus | 1970 | 879 (23.6) | 18 | 1047 (20.6) | 21 | <0.001 |
| Current Smoker | 1664 | 915 (24.5) | 24 | 697 (13.7) | 7 | <0.001 |

**Supplemental Table 2.** Association of each PRS with incident MI in participants >50 years stratified by Caribbean and Mainland subgroup.

| PRS | Analytic HR<br>(95% CI)<br>N=3682 (57 events) | Caribbean HR<br>(95% CI)<br>N = 1772 (33 events) | Mainland HR<br>(95% CI)<br>N = 1839 (22 events) |
| --- | --- | --- | --- |
| PGS000013<br>LDPred | 1.46 (1.08, 1.97) | 1.24 (0.85, 1.79) | 2.02 (1.06, 3.83) |
| PGS001355<br>AnnoPred | 1.57 (1.14, 2.15) | 1.33 (0.84, 2.08) | 1.97 (1.17, 3.30) |
| PGS002776<br>SCT | 1.21 (0.86, 1.69) | 1.01 (0.63, 1.63) | 1.70 (0.98, 2.93) |
| PGS003725<br>LDpred2 | 1.52 (1.03, 2.25) | 1.11 (0.67, 1.84) | 2.85 (1.56, 5.23) |

Note: models are adjusted for sex, first 5 principal components and is weighted for complex survey design

**Supplemental Table 3.** Associations of LDPred and AnnoPred PRS with incident MI stratified by sex and subgroup.

| PRS, Strata | Female |  |  |  | Male |  |  |  |
| --- | --- | --- | --- | --- | --- | --- | --- | --- |
|  | Sample<br>n | Incident<br>MI<br>N (%) | HR (95% CI) | C-index (SE) | Sample<br>n | Incident<br>MI<br>N (%) | Male<br>HR (95% CI) | C-index (SE) |
| AnnoPred | 4425 | 33 (0.7) | 2.39 (1.47, 3.89) | 0.844 (0.048) | 2823 | 46 (1.6) | 1.20 (0.92, 1.56) | 0.614 (0.064) |
| Caribbean<br>subset | 1185 | 21 (1.8) | 1.88 (1.15, 3.08) | 0.782 (0.064) | 1239 | 22 (1.8) | 0.98 (0.69, 1.39) | 0.611 (0.104) |
| Mainland<br>subset | 2471 | 11 (0.4) | 3.72 (1.85, 7.48) | 0.929 (0.038) | 1500 | 22 (1.5) | 1.41 (0.96, 2.06) | 0.738 (0.073) |
| LDPred | 4425 | 33 (0.7) | 2.14 (1.31, 3.49) | 0.835 (0.053) | 2823 | 46 (1.6) | 1.15 (0.89, 1.49) | 0.610 (0.068) |
| Caribbean<br>subset | 1185 | 21 (1.8) | 1.70 (1.03, 2.79) | 0.777 (0.068) | 1239 | 22 (1.8) | 0.92 (0.70, 1.22) | 0.612 (0.1030) |
| Mainland<br>subset | 2471 | 11 (0.4) | 3.55 (1.52, 8.31) | 0.917 (0.044) | 1500 | 22 (1.5) | 1.56 (0.99, 2.46) | 0.748 (0.067) |

Note: All models are adjusted for age, first 5 principal components, and weighted for complex survey design

**Supplemental Figure 1.**
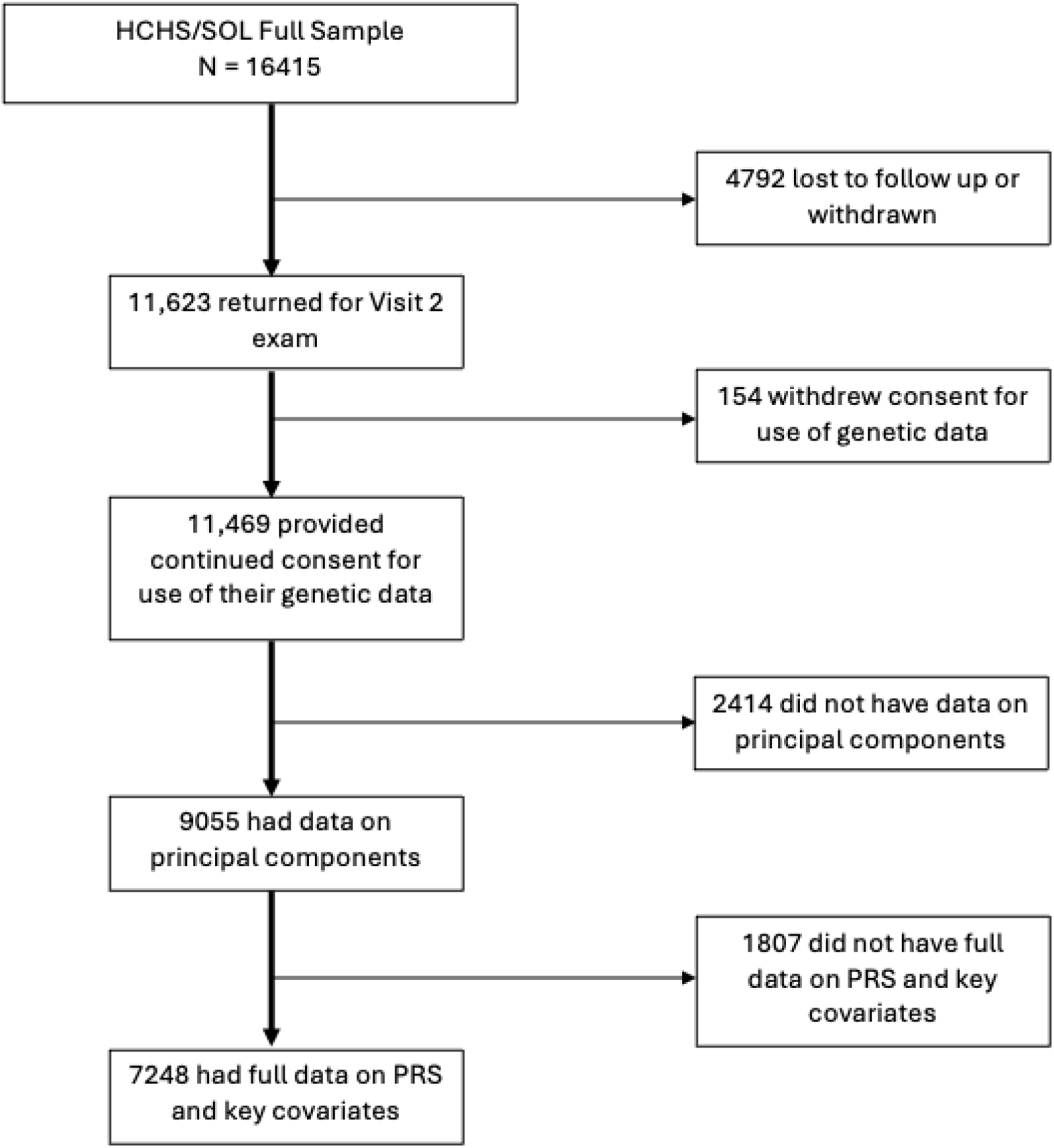

**Supplemental Figure 2.**
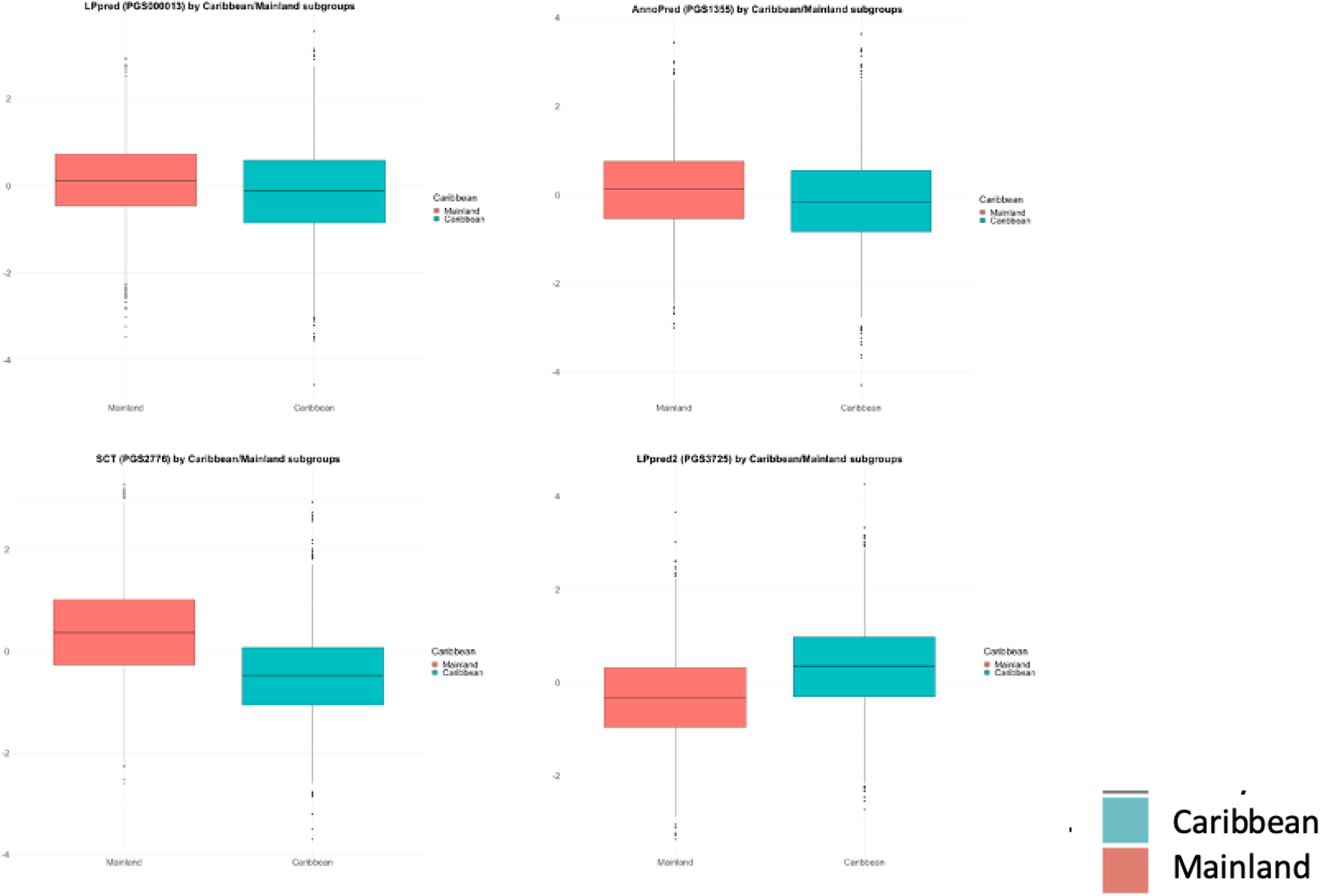
Boxplot distributions of PRS

